## Supplemental files for "Mind the gap: Mapping Variation between National and Local Clinical Practice Guidelines for Acute Paediatric Asthma from the United Kingdom and the Netherlands"

**S1 Table. Evidence and considerations outlined for second-line APA drug recommendations in sampled local British and Dutch CPGs.** Evidence grades are assigned based on a standardized BTS/SIGN methodology.

| | Intravenous MgSO4 | | | Intravenous salbutamol | | | Intravenous aminophylline | | |
|---|---|---|---|---|---|---|---|---|---|
| | Position among 2nd-line drugs | Evidence grade or mention of sources | Considerations on order of administration | Position among 2nd-line drugs | Grade of evidence or mention of sources | Considerations on order of administration | Position among 2nd-line drugs | Evidence grade or mention of sources | Considerations on order of administration |
| **BTS/ SIGN** | 1 or 2 or 3 | 1+ | Safe treatment for APA not responding to first-line drugs. Occurrence of hypotension as side effect is rare. | 1 or 2 or 3 | 1+ | One study showing that an intravenous bolus of salbutamol given in addition to near-maximal doses of nebulised salbutamol resulted in clinically significant benefits in moderate to severe asthma. | 1 or 2 or 3 | 1-<br>2+ | Side effects are common and troublesome. Evidence of benefit in children with APA unresponsive to multiple doses of B2 agonists and steroids (although loading dose was double from that which is currently recommended dose in the UK, a third of the patients withdrawn from active medication because of vomiting.) |
| **NVK** | 1 | No | None | 2 | No | None | Not | No | None outlined |
| **UK1** | 2 | No | None | 1 | No | None | 3 | No | None outlined |
| **UK2** | 1 | No | None | 2 and 4 | No | None | 3 | No | None outlined |
| **UK3** | 1 | Yes | Inexpensive, minimal adverse effects at indicated doses, dispensed on ward | 2 | | In the UK mostly dispensed in HDU. Accepted treatment option for children rather than aminophylline. | 3 | | In the UK mostly dispensed in HDU |
| **UK4** | 1 | No | Can be given anywhere in the hospital if needed acutely | 2 | No | Can be started anywhere in the hospital if needed acutely. Children receiving salbutamol infusions are usually managed on HDU. | 3 | No | None outlined |
| **UK5** | 1 | No | None | 2 | No | None | 3 | No | None outlined |
| **UK6** | 1 | No | None | 2 or 3 | No | None | 2 or 3 | No | None outlined |
| **UK7** | 1 or 2 or 3 | No | None | 1 or 2 or 3 | No | None | 1 or 2 or 3 | No | None outlined |
| **NL1a** | 1 | Yes | Side effects: hypotension, muscle weakness, drowsiness. Systematic reviews, including | 2 | Yes | The rationale for administering intravenous (as opposed to inhaled) salbutamol is likely | 3 | Yes | There is no clear evidence towards or against the use of intravenous theophylline versus intravenous salbutamol in children or |

| | | | | | | | | | |
|---|---|---|---|---|---|---|---|---|---|
| | | | some with children participants do no show a clear effect of intravenous MgSo4 when used as standard therapy for severe acute asthma. An effect has however been shown when added to standard bronchodilators and corticosteroid therapy. One paediatric study demonstrated that intravenous MgSO4 primarily reduced the need for respiratory support in the first hour of presentation. There is insufficient evidence to support the use of inhaled MgSO4. There is no literature on the effect of MgSO4 used in combination with intravenous salbutamol in children. It is also unclear whether re-administering a dose of MgSO4 is effective. Following a pragmatic approach, one may administer a second dose of MgSO4 if the first dose was associated a clear positive effect. Beware of muscle weakness as a side effect in case of near exhaustion. | | | related to the idea that inhalation therapy will not ensure therapeutic levels in case of severe airway/bronchi obstruction. However, there is insufficient evidence on whether intravenous salbutamol is safe and more effective than other therapies in children with severe acute asthma.<br><br>The usefulness of an intravenous salbutamol loading dose after frequent salbutamol inhalations is currently under discussion. At this stage, administering a salbutamol loading dose seems to be no added value. The latter effect is being explored by the Dutch STATIC-IV study.<br><br>There are no high-quality paediatric studies on the pharmacokinetics of salbutamol. It is likely that the pharmaco-kinetic properties of salbutamol are in large part similar in adults and children. From this ensues that the doses advised for use in children with APA are much higher than the recommended doses in adults. The safety and effectiveness of the frequent use of high doses of intravenous salbutamol in children with severe acute asthma thus appears highly questionable. Based on the literature it is advisable to avoid a prolonged use of high doses of intravenous salbutamol in children. | | | adults. Xanthine derivates may cause more side effects. A recent RCT found a longer length of stay for children having received an adequate dose of aminophylline intravenous rather than no aminophylline or an inadequate dose of aminophylline. Inhaled aminophylline does not improve patient outcomes. Adding aminophylline or theophylline to the standard bronchoinhalation therapy for APA does not seem to be effective. While there is possibly a short-term effect on lung function, no clear evidence has been found on hard outfcomes such asthma scores, length of admission, the need for intubation etc. The costs of intravenous aminophylline are significantly lower than salbutamol intravenous however. Based on the curreent literature, the side effects, we have chosen to only advise theophyllin as an alternative therapy.. |
| **NL1b** | 1 | No | None | 2 | No | None | Not recommended | | |

| | | | | | | | | | |
|---|---|---|---|---|---|---|---|---|---|
| **NL2** | 1 | | No large-scale studies | 2 | Yes | No evidence for theophylline versus salbutamol intavenous in adults | 3 | Yes | No evidence for theophylline compared to salbutamol intavenous in adults but theophylline has a small therapeutic range and is a xanthine derivate which gives more side effects (*listed in CPG*), hence the use of theophylline only to prevent intubation. |
| **NL3** | 1 | No | None | 2 | No | None outlined | Not recommended | | |
| **NL4** | 1 | No | Does not need to be administered in high care setting. | 2 | No | Can be started in A&E before child is transported to PICU. | Not recommended. | | |
| **NL5** | 1 | No | None | 2 | No | Administer in intensive care | Not recommended. | | |
| **NL6** | 1 | No | None | 2 | No | Can be started before child is transported to ICU | Not recommended. | | |
| **NL7** | 1 | Yes | Positive effects on severe asthma, no relevant side effects. Safe and effective in adults according to meta-analysis. Convenient administration, no subsequent delays. | 2 | Yes | None outlined. | Not recom-mended | Yes | Significantly smaller therapeutic range than beta2-agonists. Recommends to be cautious with toxicity. Cites two studies finding no added value alongside therapy with corticosteroids and beta2-agonists. |
